# Forecasting seizure likelihood with wearable technology

**DOI:** 10.1101/2021.05.20.21257495

**Authors:** Rachel E. Stirling, David B. Grayden, Wendyl D’Souza, Mark J. Cook, Ewan Nurse, Dean R. Freestone, Daniel E. Payne, Benjamin H. Brinkmann, Tal Pal Attia, Pedro F. Viana, Mark P. Richardson, Philippa J. Karoly

**Affiliations:** Department of Biomedical Engineering, The University of Melbourne; Departments of Medicine and Neurology, The University of Melbourne, St Vincent’s Hospital, Melbourne; Graeme Clark Institute for Biomedical Engineering, The University of Melbourne; Seer Medical, Australia; Bioelectronics Neurophysiology and Engineering Lab, Department of Neurology, Mayo Clinic, Rochester, MN; School of Neuroscience, Institute of Psychiatry, Psychology and Neuroscience, King’s College London, London, UK; Faculty of Medicine, University of Lisbon, Portugal

## Abstract

The unpredictability of epileptic seizures exposes people with epilepsy to potential physical harm, restricts day-to-day activities, and impacts mental well-being. Accurate seizure forecasters would reduce the uncertainty associated with seizures but need to be feasible and accessible in the long-term. Wearable devices are perfect candidates to develop non-invasive, accessible forecasts but are yet to be investigated in long-term studies. We hypothesized that machine learning models could utilize heart rate as a biomarker for well-established cycles of seizures and epileptic activity, in addition to other wearable signals, to forecast high and low risk seizure periods.

This feasibility study tracked participants’ (n = 11) heart rates, sleep, and step counts using wearable smartwatches and seizure occurrence using mobile seizure diaries for at least 6 months (mean = 14.6 months, SD = 3.8 months). Eligible participants had a diagnosis of refractory epilepsy and reported at least 20 seizures (mean = 135, SD = 123) during the recording period. An ensembled machine learning and neural network model estimated seizure risk either daily or hourly, with retraining occurring on a weekly basis as additional data was collected. Performance was evaluated retrospectively against a rate-matched random forecast using the area under the receiver operating curve. A pseudo-prospective evaluation was also conducted on a held-out dataset.

Of the 11 participants, seizures were predicted above chance in all (100%) participants using an hourly forecast and in ten (91%) participants using a daily forecast. The average time spent in high risk (prediction time) before a seizure occurred was 37 minutes in the hourly forecast and 3 days in the daily forecast. Cyclic features added the most predictive value to the forecasts, particularly circadian and multiday heart rate cycles.

Wearable devices can be used to produce patient-specific seizure forecasts, particularly when biomarkers of seizure and epileptic activity cycles are utilized.

## Introduction

Epilepsy is one of the most common neurological disorders, affecting roughly 1% of the world’s population (1) and responsible for 20.6 million disability-adjusted life-years (DALYs) lost, which is comparable to breast cancer in women and lung cancer in men (2). Epilepsy is characterized by an increased predisposition of the brain to generate epileptic seizures, which often result in vast neurobiological, cognitive, psychologic, and social consequences (3). Despite decades of new drug development and surgical treatment, up to one-third of people with epilepsy continue to suffer from recurrent seizures (4,5). While most people are symptom-free for more than 99.9% of their day-to-day life, epileptic seizures are sudden, potentially catastrophic events that can be life-threatening both for the person with epilepsy and others. Crucially, sudden death in epilepsy (SUDEP), most often following a convulsive seizure, is 27 times more likely than sudden death in control populations, a mortality burden second only to stroke when compared to other neurologic diseases (6,7). Aside from these risks, living with epilepsy can take a major toll on quality of life and independence, as the unpredictable nature of seizures causes feelings of uncertainty (8) and impacts participation in common day-to-day activities, such as going to work, driving, and social interactions (9).

To address the uncertainty associated with epileptic seizures, researchers across many disciplines have spent years investigating the potential for seizure prediction and forecasting (10). The ability to reduce the uncertainty of when a seizure is about to occur would have tremendous implications for quality of life, and clinical management (10). Timely precautions against seizure-related injury or timed adjustment of treatment according to seizure likelihood (chronotherapy) could also reduce seizure-related harm, hospitalizations, and healthcare-related costs (11).

Until recently, there was no scientific consensus as to whether seizures would be predictable in a prospective setting since most research was based on limited data (from short-duration in-hospital electroencephalography (EEG) recordings) and some presented methodological flaws (12). Access to better quality data (made available in public databases (13,14) and seizure prediction competitions (15)), more rigorous statistical and analytical methods, and results from a clinical trial of an intracranial EEG seizure advisory system (NeuroVista (16)) have shown promise that seizure prediction devices could be possible in the foreseeable future. Additionally, there is a better understanding of the pre-seizure state and of the mechanisms underlying seizure generation (ictogenesis), with contributions from basic science, network theory, multiscale electrophysiological recordings, and functional neuroimaging (17). Multiple patient-specific seizure precipitants have also been identified, including stress (18,19), poor sleep (18), exercise (20), diet (21), weather (22,23), alcohol use (24) and poor drug adherence (25). Many of these factors have shown potential utility in forecasting seizures (18,23).

Yet perhaps the most significant breakthrough for the field of seizure forecasting has been the characterization of short- and long-term seizure occurrence cycles (11,26,27), which typically occur in circadian and multiday (often weekly and monthly) periodicities (27,28). Similar cycles have been reported in interictal epileptiform activity (IEA) (26), EEG markers of brain critical slowing (29) and heart rate (30), all of which have been linked to seizure timing, suggesting that seizures are co-modulated by underlying biological cycles. An individual’s seizure cycles can be utilized to generate seizure forecasts using both self-reporting seizure diaries (31–33) and electrographic seizures (34). However, the discrete nature of seizure events means that the underlying biological cycles may be stronger predictors of seizure occurrence than seizure cycles alone (29,34,35). This has already been successfully demonstrated with cycles of IEA in a retrospective seizure forecasting study using an implanted intracranial EEG device (34). Furthermore, algorithms incorporating biological cycles seem to outperform algorithms using more traditional EEG features, such as spectral power and correlation (15).

However, seizure forecasting algorithms typically rely on chronic EEG recordings from invasive, implanted devices, which require surgery (and associated risks), are costly, and may not be an option for many people with epilepsy. Minimally-invasive or noninvasive wearable devices that monitor continuous biomarkers of seizure risk are, therefore, ideal candidates for most people who desire seizure forecasts (9). Currently, some wearable devices are commercially available for seizure *detection* (36), although there are also promising results highlighting the utility of wearables in seizure forecasting. Wearable sensors can be used to detect actigraphy, blood volume pulse, body temperature, cerebral oxygen saturation, electrodermal activity and heart rate, all of which have all shown promise in seizure prediction (37–39). Periodic wearable signals, such as temperature (40) and heart rate (30) may also be used as a biomarker for seizure cycles (35). For example, our recent work in seizure timing and heart rate, measured from a wearable smartwatch, shows that seizures are often phase-locked to underlying circadian and multiday cycles in heart rate (i.e., there is a strong preference for seizures to occur at specific phases of the heart rate cycle) (30).

To address the need for non-invasive seizure forecasting, this study aimed to develop a wearable device-based seizure forecaster using a long-term dataset from an observational cohort study, Tracking Seizure Cycles. We hypothesized that cycles in heart rate can be leveraged, in addition to other wearable signals (other heart rate features, step count and sleep features), to forecast high and low seizure risk periods. We also investigated the relative contributions of cycles, heart rate, sleep and activity features to forecasting performance.

## Materials and Methods

### Study design and participants

This retrospective and pseudo-prospective feasibility study was designed using training and testing datasets, followed by pseudo-prospective evaluation using a held-out dataset. We utilized long-term mobile seizure diaries and a wearable smartwatch to forecast seizure likelihood and elucidate the relationship between seizures and non-invasively measured wearable signals, namely heart rate, sleep stages, sleep time, and step count. Seizures are known to follow circadian and multiday cycles in most people, so there was a specific focus on finding physiological cycles with similar periodicities using wearable signals. This builds on our previous work that found a relationship between cycles in heart rate and seizure occurrence (30). The study was approved by the St Vincent’s Hospital Human Research Ethics Committee (HREC 009.19) and all participants provided written informed consent. First enrollment was in August 2019.

All participants were from the observational cohort study ‘Tracking Seizure Cycles’ (TSC). Participants were over 18 years of age with a confirmed epilepsy diagnosis, and either uncontrolled or partially controlled seizures. We chose to recruit an unselected cohort, as seizure cycles are known to be present regardless of age, epilepsy syndrome, and seizure type and frequency (26,27). Continuous data were collected via mobile and wearable devices for at least 6 months and up to 20 months. Participants wore a smartwatch (Fitbit, Fitbit Inc., USA) and manually reported seizure times in a freely available mobile diary app (Seer App, Seer Medical Pty Ltd, Australia). The smartwatch continuously measured participants’ heart rates (via photoplethysmography) at 5 s resolution (one recording every 5 seconds). The smartwatch also estimated sleep stage (awake, REM, and light and deep sleep) and step count each minute.

Participants were required to have two months or more of continuous wearable data recordings, at least 80% adherence (i.e., they must have worn the device at least 80% of the time) and a minimum of 20 seizures reported during the recording time to be eligible for seizure forecasting. Eligible participant demographic information is given in Table 1.

**Table 1.** Eligible participants’ demographic information. Participants that had more than one seizure during the evaluation period are shown in red.

| Participant | Type of seizures (Focal, Generalized or Both) | Total seizures during monitoring (frequency/month) | Training recording length (months) | Testing recording length (months) | Evaluation recording length (months) | Sleep scoring (nights) |
| --- | --- | --- | --- | --- | --- | --- |
| P1 | Focal | 57 (5.1) | 4.2 | 4.3 | 2.7 | 334 |
| P2 | Focal | 111 (8.8) | 2.0 | 7.9 | 2.7 | 371 |
| P3 | Focal | 27 (1.5) | 12.6 | 3.1 | 2.7 | 549 |
| P4 | Focal | 24 (1.4) | 10.6 | 4.3 | 2.7 | 500 |
| P5 | Both | 280 (17.0) | 2.0 | 13.3 | 1.2 | 459 |
| P6 | Focal | 246 (36.7) | 2.0 | 2.1 | 2.6 | 199 |
| P7 | Generalized | 28 (1.6) | 8.5 | 5.9 | 2.7 | 501 |
| P8 | Focal | 179 (14.6) | 2.5 | 7.1 | 2.7 | 327 |
| P9 | Both | 392 (19.6) | 9.7 | 7.6 | 2.7 | 586 |
| P10 | Focal | 94 (6.6) | 2.4 | 9.2 | 2.7 | 428 |
| P11 | Focal | 55 (3.9) | 3.5 | 7.8 | 2.7 | 399 |
| Summary | 8 focal only, 1 generalised only, 2 both generalised and focal | M = 136 (10.6)<br>SD = 123 (10.8) | M = 5.5<br>SD = 4.0 | M = 6.7<br>SD = 3.2 | M = 2.6<br>SD = 0.5 | M = 423<br>SD = 112 |

The seizure forecast was presented in hourly and daily formats to assess the accuracy of an hourly forecast compared to a daily forecast. The hourly forecast gave the likelihood of a seizure at the start of the hour, every hour. The daily forecast gave the likelihood of a seizure for the day, shortly after waking from sleep (based on Fitbit’s sleep end time).

### Training, testing and held-out evaluation datasets

The training dataset included at least 2 months of continuous recordings (M = 5.4 months, SD = 4 months) and at least 15 seizures (M = 35, SD = 47). The patient-specific training cut-off date was the final day that both of these criteria were met. The testing dataset included participants’ continuous recordings (M = 6.6 months, SD = 3.1 months) and seizures (M = 87, SD = 112) reported from their training cut-off date until 1 February 2021. As a further requirement for seizure forecasting, participants must have had at least five lead seizures (at least an hour apart in the hourly forecast and at least a day apart in the daily forecast) reported during the testing period. Any continuous recordings (M = 2.6 months, SD = 0.5) and seizures (M = 13, SD = 14) reported from 1 February 2021 until 25 April 2021 were included in the held-out evaluation cohort, so long as the participant reported at least one seizure during this period. This data was held-out to evaluate the efficacy of the forecasting algorithm in a pseudo-prospective setting.

### Data preprocessing

The heart rate, step count, and sleep signals were all processed separately. Heart rate features included rate of change in heart rate (RCH) and daily resting heart rate (RHR). Physical activity features included steps recorded in the previous hour and steps recorded on the previous day. Sleep features included total time asleep (not including naps), time in REM, time in deep and light sleep during main sleep, average HR overnight, sleep time deviation from median sleep time over the past three months, and wake time deviation from median wake time over the past three months. All sleep features were calculated using sleep labels derived from Fitbit’s sleep algorithm. Additionally, we included cyclic features, comprising heart rate cycles (circadian and multiday), last seizure time, and second-last seizure time. Compared to the hourly forecast, the daily forecast only included multiday cycles, days since last seizure time, days since second-last seizure time, all sleep features, daily resting heart rate, and steps recorded during the previous day.

To derive heart rate features and heart rate cycles, continuous heart rate signals were initially down-sampled to one timestamp per minute, followed by interpolation of short missing data segments with a linear line (missing segments less than two hours) or longer missing data segments with a straight line at the mean heart rate. RCH was used to estimate heart rate variability (HRV), which is defined as the variations in RR intervals and is typically derived using the QRS complex on an electrocardiogram (ECG). RCH was calculated as the mean beats per minute (BPM) in one minute subtracted from the mean BPM in the previous minute, representing the change in BPM over two minutes. RCH was resampled every hour for the hourly forecast or every day for the daily forecast. Daily RHR was derived as the average of the bottom quintile of BPM where no steps were recorded.

To compute the heart rate cycles, we used a similar approach to a method used to extract multiday rhythms of epileptic activity (26) (see also (30) for further details). Briefly, circadian and multiday peak periodicities of heart rate (cycles) were derived using a Morlet wavelet. The heart rate signal was filtered (using a zero-order Butterworth bandpass filter) at the peak periodicities and instantaneous phase of the cycle at each timepoint was estimated using a Hilbert transform. Cycles were used as features for the forecaster if seizures were significantly phase-locked to the cycle (p<0.05, according to Omnibus/Hodges-Ajne test for circular uniformity (41)). Each cyclic feature (cycle phases and last/second last seizure time) was transformed into two linear features by normalizing the signal from 0 to 2π and computing the sine and cosine.

### Forecasting algorithm

Hourly and daily forecasts were generated for each participant. To forecast the likelihood of a seizure hourly and daily, we used an ensemble of a long short-term memory (LSTM) neural network (42), a random forest (RF) regressor (42), and a logistic regression (LR) classifier (42). Figure 1 describes the architecture of the model. The training (green), testing (orange) and evaluation (red) cohorts were different lengths in each participant, and algorithm retraining occurred weekly during testing and evaluation. The model used sleep features in the LSTM model (which contains 7 days of memory) in order to account for the potential effect of built-up sleep debt on seizure risk (18). All other features (cycles, heart rate features and step counts) then predicted seizure risk using a random forest model, which performed best by observation. A logistic regression model, which weighs inputs’ predictive value, then combined the random forest and LSTM outputs into one seizure risk value per hour or day. This was compared to a rate-matched random model (occasionally referred to as the chance model) using AUC scores. Other metrics were also used to assess forecast performance (see *Performance metrics*).

**Figure 1.**
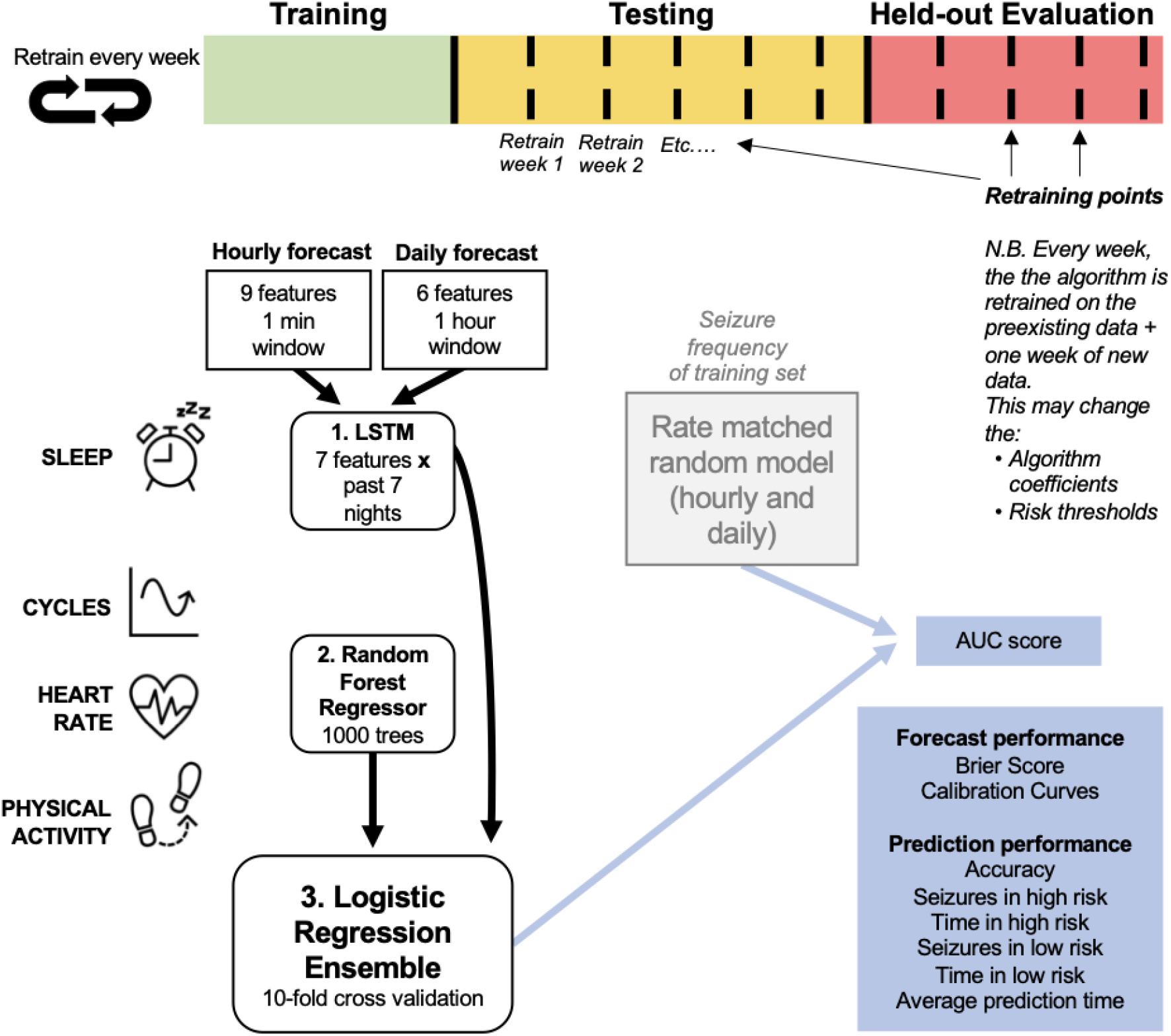
Forecasting model architecture. The logistic regression ensemble (combining LSTM, Random Forest Regressor, and all features) was trained on a training dataset that included at least 15 seizures and at least two months of continuous recordings. Two forecasting horizons were compared: hourly and daily forecasts. The LSTM model incorporated sleep features from the past seven nights and the random regressor included all other features (cycles, heart rate, and physical activity features), in addition to the output daily seizure likelihood estimates from the LSTM model. The logistic regression ensemble utilized a 10-fold cross validation approach to forecast seizure likelihood hourly or daily. The forecasting model was assessed (using AUC scores) on a retrospective testing set and a pseudo-prospective held-out evaluation set and compared to a rate-matched random (RMR) model, where seizure frequency was determined by the training set. The algorithm was retrained weekly to imitate a clinical forecast.

The LSTM model was trained on sleep features computed daily after waking. A weekly history of sleep features was incorporated into each row input, providing a 7×7 matrix for each forecast, representing 7 days and 7 sleep features per day. The LSTM model was composed of a single layer with 64 memory units, followed by two densely connected layers, and a linear activation function. All networks were trained for 100 epochs. We selected the mean squared error loss function as the cost function, using the Adaptive Moment Estimation (Adam) optimizer (43). The LSTM model outputted the likelihood of a seizure for the day based on sleep features and was used as an input to the LR classifier.

The RF regressors with the bootstrap aggregating technique were trained on all physical activity, heart rate, and cyclic features. In the model, the number of decision trees was 1000 and the minimum number of samples required to be at a leaf node was 120. From observation, these model parameters achieved the highest accuracy across the board during training. Most people, particularly participants with low seizure frequency (<2 seizures/month), had a highly imbalanced dataset, with non-seizure hours/days occurring far more frequently than seizure hours/days. RF models typically performed better on balanced datasets, so oversampling of seizure hours/days was undertaken before training the RF model. The output of the RF model was the likelihood of a seizure within the following hour or day and was used as an input to the LR classifier.

The LR classifiers were trained on the outputs of the LSTM and RF models. To aid the classifier in distinguishing between non-seizure hours/days and seizure hours/days and remove the impact of pre-ictal and post-ictal changes, the hour/day immediately preceding and following the hour/day of each seizure were removed in the training dataset. The output of the LR model was the final likelihood of a seizure (risk value); the risk value was represented as a continuous value between 0 for no seizure and 1 for a ‘guaranteed’ seizure within the next hour or day, as appropriate.

The forecaster classified hours and days as either low, medium, or high risk. The medium and high risk cut-off thresholds were computed using the training dataset by optimizing the metrics:

(C1) time spent in low risk > time spent in medium risk > time spent in high risk;
(C2) seizures in high risk > seizures in medium risk > seizures in low risk (29).
If C1 or C2 could not be satisfied, the optimization algorithm maximized the product of the time in low risk and the number of seizures in high risk (C3 and C4):
(C3) maximize the time spent in a low risk state;
(C4) maximize the number of seizures occurring in the high risk state.

Retraining the algorithm was implemented to imitate a clinical seizure forecasting device in which algorithm coefficients and risk thresholds would be regularly updated. Retraining of the seizure forecast occurred on a weekly basis as additional data was collected.

### Performance metrics

To assess the performance of the hourly and daily forecasters, a variety of different metrics were used. During algorithm testing and for pseudo-prospective held-out evaluation, performance of the ensembled model was evaluated using the area under the receiver operating characteristic curve (AUC) and compared to the AUC score of a rate-matched (seizure frequency derived from all seizures that occurred in the training dataset) random forecast. The AUC scores assessed the classifier’s ability to discriminate between non-seizure hours/days and seizure hours/days.

Despite the usefulness of the AUC to measure performance, the AUC can change depending on the forecasting horizon (34); in this case, an hourly forecast compared to a daily forecast. This motivated the use of Calibration Curves (CC) to measure how well the predicted likelihood values corresponded to observed probabilities, and the Brier score (or Brier loss) to quantify the accuracy of the predictions. The CC metric provides a visual representation of the forecaster’s ability to estimate seizure risk. The ideal CC can be visualized as a diagonal line, where the forecaster’s predicted seizure likelihood values are equal to the actual seizure probabilities. Anything above this line would be considered underestimating seizure risk and anything below would be overestimating seizure risk. The Brier score (or Brier Loss) is shown alongside the CC metric, which is often used to assess calibration performance. For the Brier Score, a perfectly accurate forecast would result in a loss score of 0 and a poorly performing forecast would result in a loss closer to 1. We also considered the accuracy of the forecaster, time spent in low, medium and high risk states, and seizures occurring in low, medium and high risk states.

Analyses were executed using Python (version 3.7.9).

## Results

There were 11 out of 39 participants that met the inclusion requirements (see *Methods: Study Design and Participants*) (Table 1). Eligible participants had an average duration of 14.6 months (SD = 3.8) of continuous heart rate and activity monitoring, and an average of 423 nights (SD = 112) that recorded sleep stages and duration. Participant diaries included an average of 136 (SD = 123) seizures reported during the wearable monitoring period. Results from the cohort are given in Figures 2-6 and Table 2. Eight of 11 participants (shown in red in Table 1) in the testing cohort were also included in the held-out evaluation cohort, as these people reported more than one seizure during the evaluation period. The results from the prospective evaluation cohort are shown in Figure 7 and Table 2.

**Table 2.** AUC scores of the hourly and daily forecasters for the testing and evaluation cohorts. ^*^ indicates performance greater than chance (the rate-matched random forecast).

|  |  | Testing dataset |  | Evaluation dataset |  |
| --- | --- | --- | --- | --- | --- |
|  |  | Hourly AUC | Daily AUC | Hourly AUC | Daily AUC |
| Participant | 1 | 0.79* | 0.64* | 0.68* | 0.42 |
|  | 2 | 0.71* | 0.61* |  |  |
|  | 3 | 0.93* | 0.62* | 0.94* | 0.69* |
|  | 4 | 0.89* | 0.92* |  |  |
|  | 5 | 0.75* | 0.72* |  |  |
|  | 6 | 0.57* | 0.46 | 0.55* | 0.62* |
|  | 7 | 0.67* | 0.64* | 0.41 | 0.45 |
|  | 8 | 0.70* | 0.70* | 0.82* | 0.80* |
|  | 9 | 0.76* | 0.68* | 0.57* | 0.45 |
|  | 10 | 0.66* | 0.66* | 0.61* | 0.80* |
|  | 11 | 0.69* | 0.61* | 0.84* | 0.45 |
| Mean (SD) |  | **0.74 (0.1)** | **0.66 (0.11)** | **0.68 (0.18)** | **0.59 (0.16)** |

**Figure 2.**
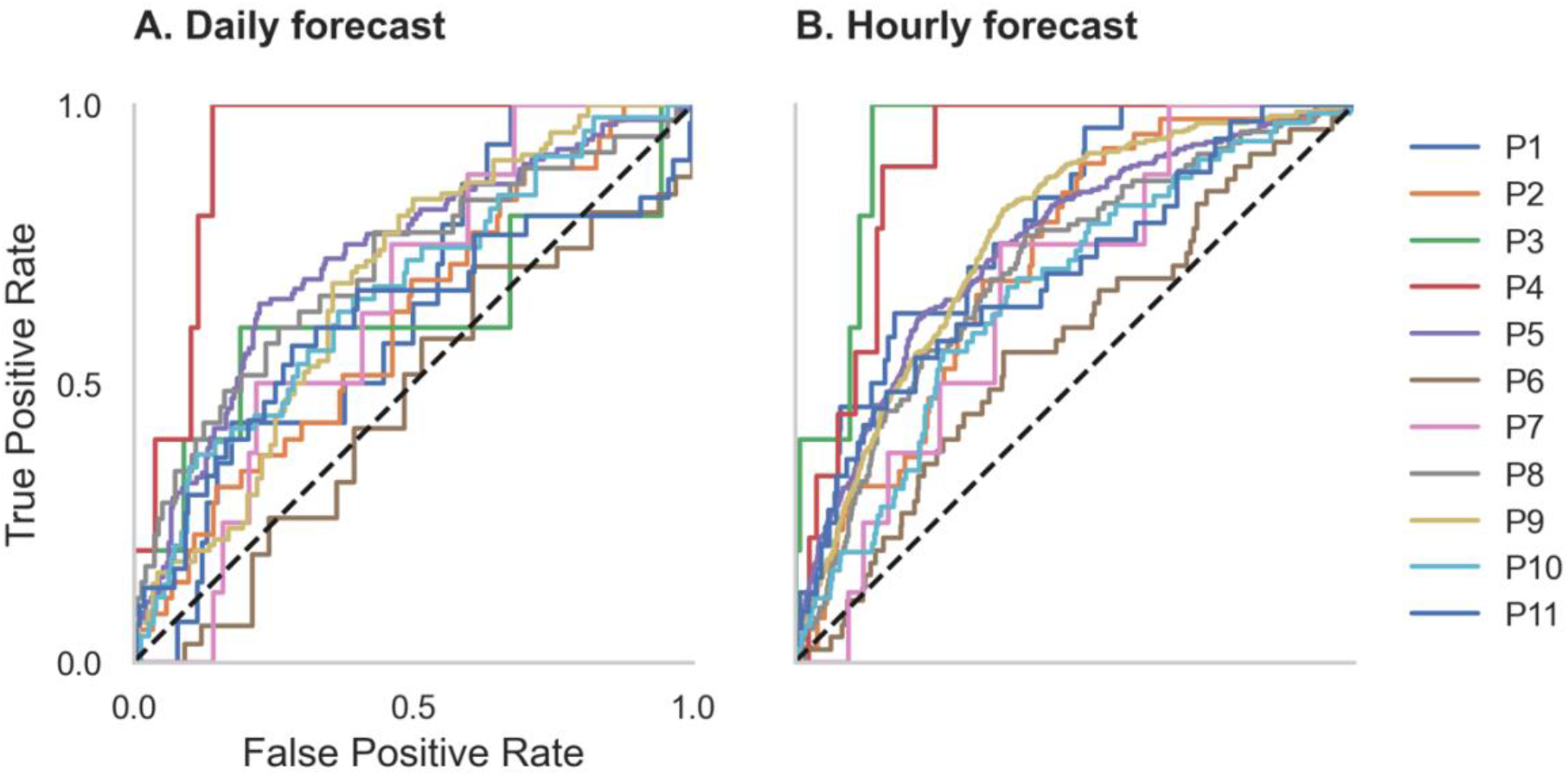
Receiver operator characteristic (ROC) curves for all participants in the daily and hourly forecast (retrospective testing cohort). The dashed diagonal line represents a balanced random forecast. ROC curves show that hourly forecasts consistently outperformed a balanced random forecaster, and daily forecasts mostly outperformed a balanced random forecaster. Patient-specific forecast performance was assessed by comparing the forecaster’s area under the ROC curve (AUC) to the AUC of a rate-matched random forecast (different to the balanced random forecast shown above).

### Forecast performance and metrics

Forecasting performance was quantified to determine which participants would have benefitted from the non-invasive seizure forecast. First, we used the AUC metric to determine forecasting performance. The AUC score quantifies how useful the forecast is, based on the amount of time spent in a high-risk state. An excellent forecast is often considered to have an AUC of greater than 0.9. Of the 11 participants, AUC scores showed that seizures were predicted above chance in all participants using an hourly forecast (M AUC = 0.74, SD = 0.10) and in 10 participants using a daily forecast (M AUC = 0.66, SD = 0.11) (Figure 2 and Table 2).

People with longer recording times usually performed well in both the hourly and daily forecasts. A weak positive correlation exited between total recording length and AUC scores in both the hourly (R^2^ = 0.63) and daily (R^2^= 0.59) forecasters (Supplementary Figure 1). This suggests that the forecaster improves over time.

A relationship was also noticed between seizure frequency and forecasting performance. The participant with the highest seizure frequency (P6) had the worst performance in both the hourly and daily forecasters (0.57 and 0.46, respectively). P6 had a seizure frequency of 36.7 seizures/month (i.e., more than one per day), which was almost double the next highest participant. Across the whole cohort, a weak negative correlation exited between seizure frequency and AUC scores in both the hourly (R^2^ = -0.58) and daily (R^2^= -0.49) forecasters (Supplementary Figure 2). This suggests that participants with lower seizure frequencies (less than once per day) perform better than participants with higher seizure frequencies.

Time spent in high, medium, and low risk, alongside the seizure frequency in high, medium, and low risk, were also considered (Figure 3). For the hourly forecast, median forecast accuracy was 86% (min: 56%, max: 95%) and median time in high risk was 14% (min: 5%, max: 45%). For the daily forecast, median forecast accuracy was 83% (min: 43%, max: 97%) and median time in high risk was 18% (min: 6%, max: 29%). Of the 11 participants, the average time spent in high risk (prediction time) before a seizure occurred was 37 minutes in the hourly forecast and 3 days in the daily forecast. Typically, greater AUC scores implied that the participant spent more time in low risk and most seizures occurred in high risk. For example, P4 spent only 7% of their time in high risk state, but 83% of their seizures occurred whilst in high risk (see Figure 5 for an example forecast).

**Figure 3.**
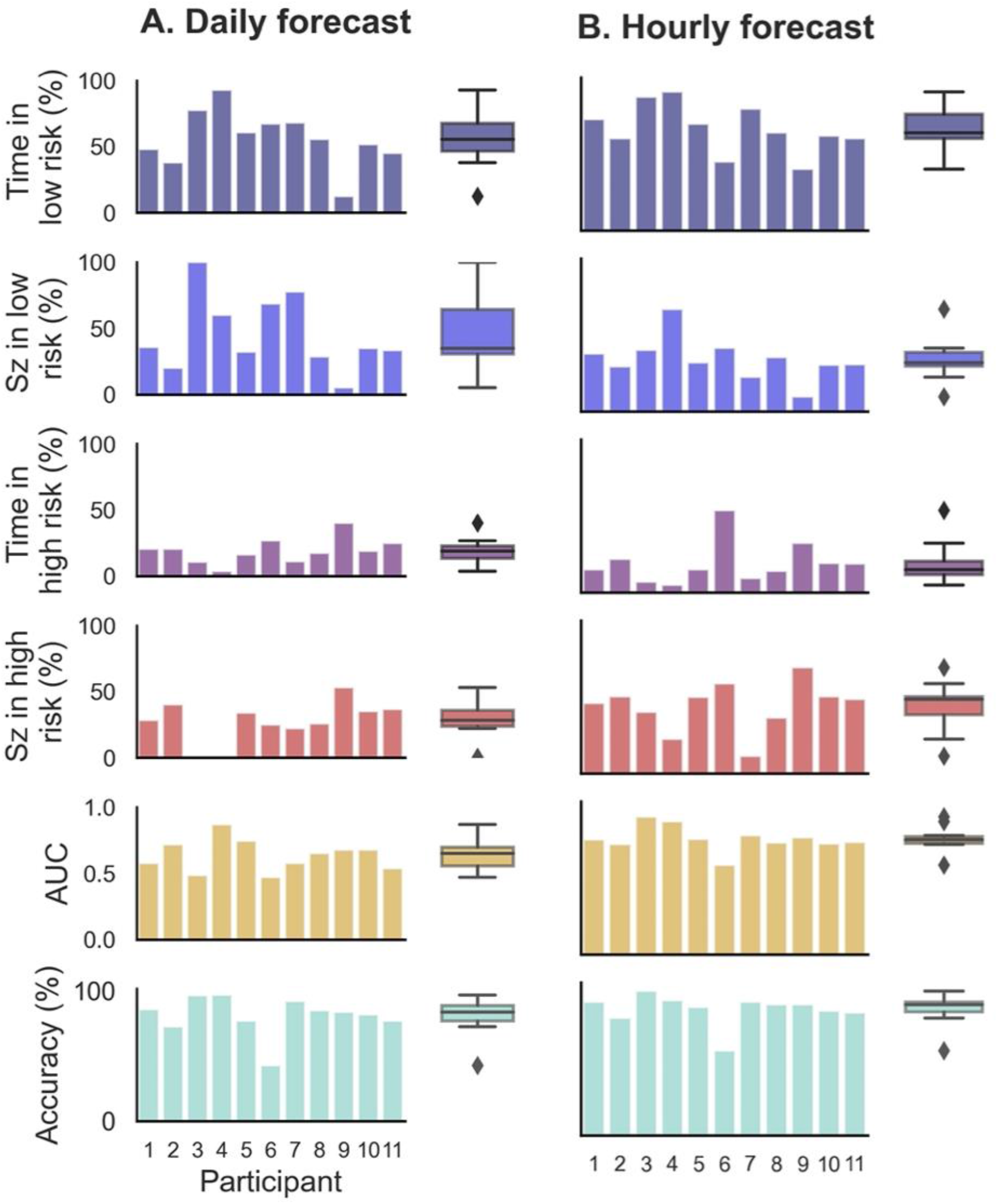
Forecasting and prediction performance metric results in the retrospective testing cohort for the hourly and daily forecasters. Individual participant bars are shown for each metric. Population box plots are shown on the right of the bars, showing median and upper and lower quartiles for each metric in the hourly and daily forecasters.

Additionally, we evaluated CC metrics and Brier scores (Figure 4). Generally, people with more seizures had calibration curves closer to the ideal diagonal line. Hourly and daily forecasts were occasionally found to sit well below the ideal line, suggesting that seizure risk was overestimated in these cases. Brier score loss, another metric to assess forecast calibration performance, varied independently to calibration curve variation. For example, the participants with the highest seizure counts (P5 and P9) had similar calibration curves for both the hourly and the daily forecast; however, Brier loss scores were much greater for P9 than P5. P4 had the lowest Brier loss scores in both the hourly and daily forecast.

**Figure 4.**
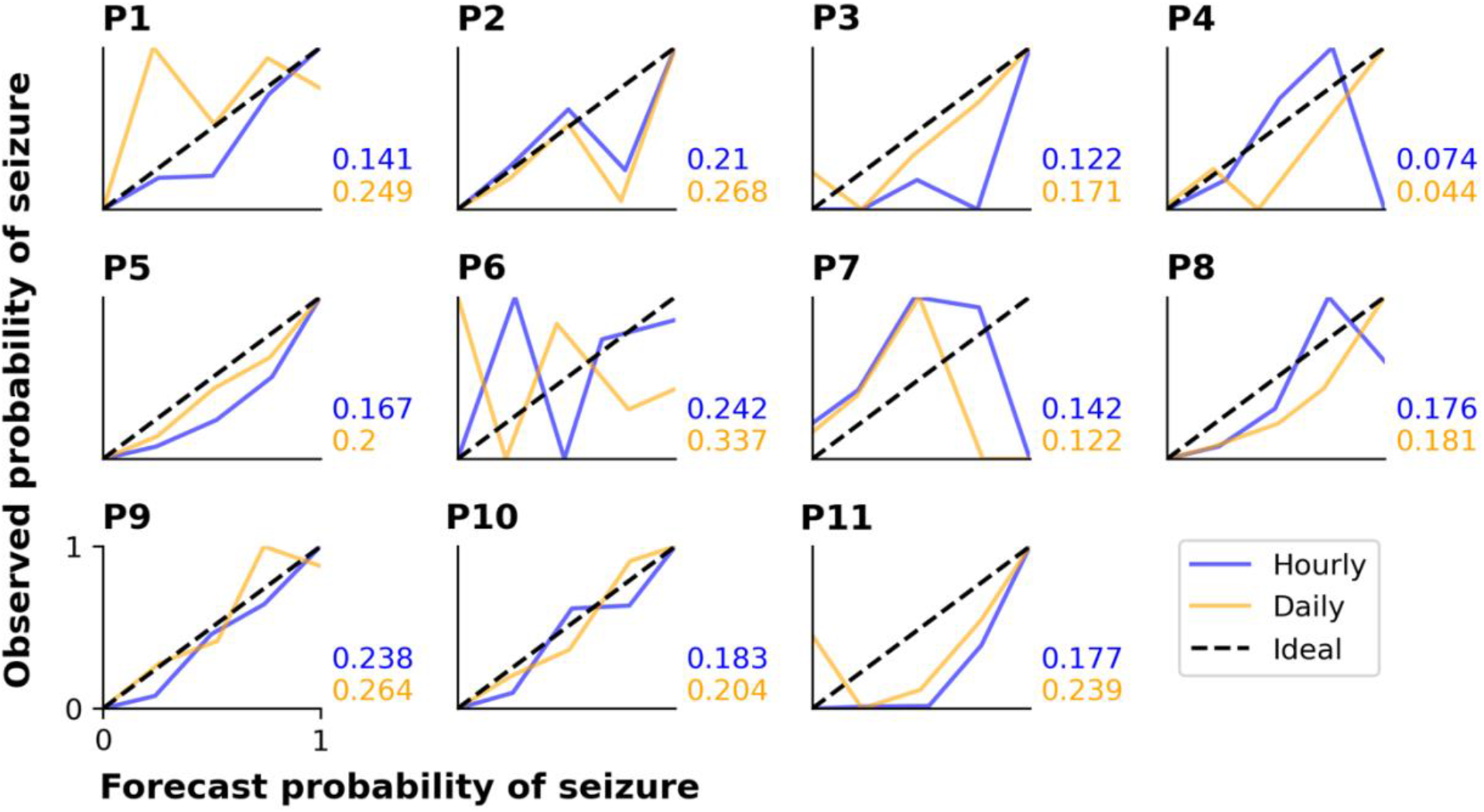
Calibration curves and Brier scores for hourly and daily forecasts summarized for each participant in the retrospective testing cohort. The calibration curves show the relationship between the forecasted likelihood of seizures and the actual observed probability of seizures. For the calibration curves, 10 bin sizes were used, so forecast likelihood values were compared to actual probabilities from 0-10%,10-20%,…,90-100%. The ideal calibration curve for a hypothetically perfect forecaster is shown in each plot.

**Figure 5.**
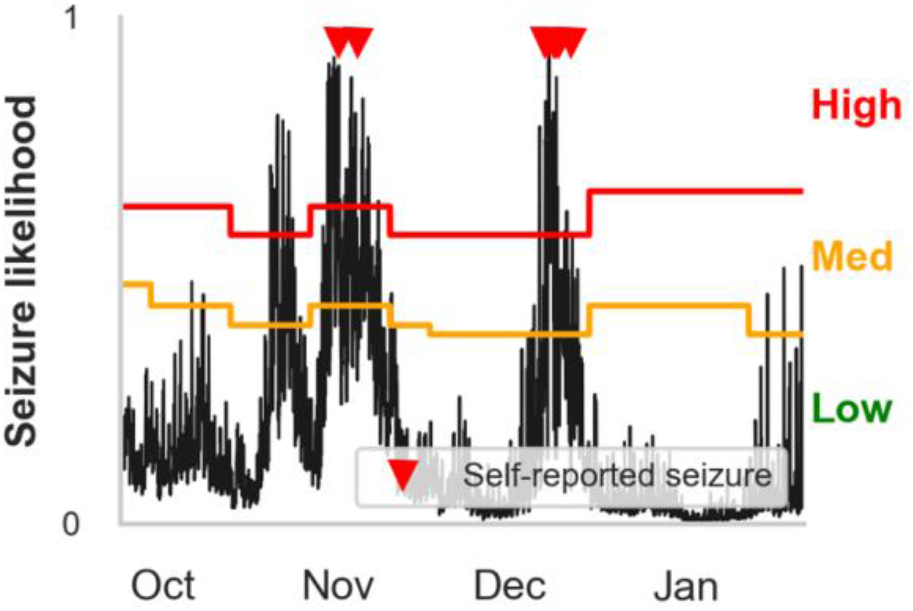
Example hourly forecasts showing high, medium, and low risk states and thresholds. Predicted seizure likelihood (black line) derived from the hourly forecaster for P4 from the end of September to the end of January. Seizures are marked with red triangles. Note that the medium risk and high risk thresholds – indicated by the orange and red lines, respectively – can change after weekly retraining. The cyclical seizure likelihood is mostly attributable to multiday heart rate cycles.

### Feature groups on forecast performance

To characterize the importance of feature groups on forecasting performance, we analyzed AUC score change with the addition of particular feature groups (Figure 6). Physical activity and heart rate feature groups added little predictive value to the daily forecaster. Sleep features added value to the daily forecaster in some people. Physical activity and heart rate features added some predictive value to the hourly forecaster; however, sleep features were the weakest predictors in the hourly forecaster. In both the hourly and daily forecaster, the cycles feature group was the strongest predictor for most people. 10 of 11 participants (all expect P4) had a significant (i.e., seizures were significantly locked onto the cycle in the training dataset) circadian cycle and 10 of 11 (all except P7) people had least one significant multiday cycle. Despite the occasional negative AUC score change with the addition of a feature group, it is important to note that it is unlikely that there is significant positive or negative value added to the forecaster when values are close to 0.

**Figure 6.**
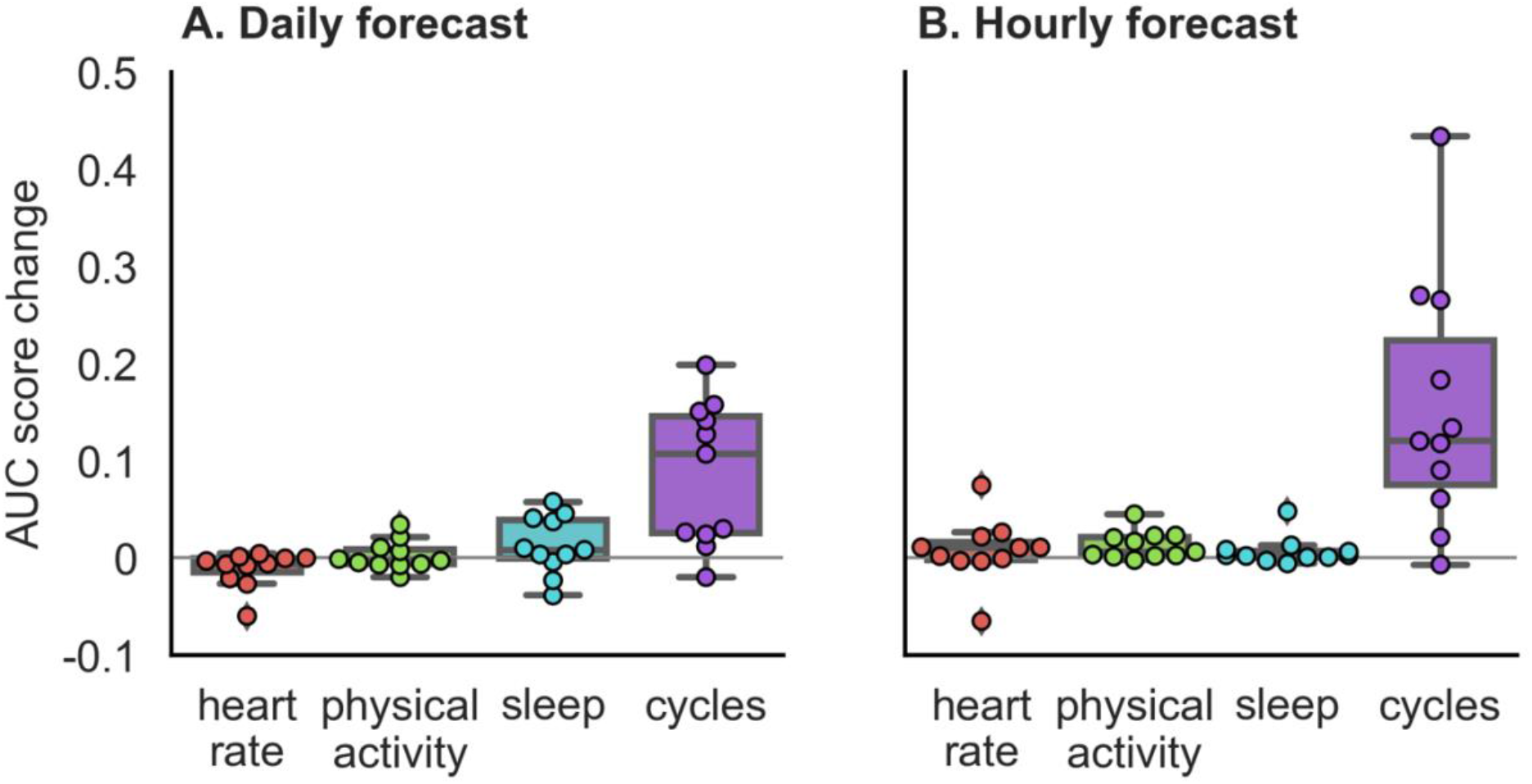
Auxiliary contribution of each feature group on forecasting performance in the retrospective testing cohort. AUC score change represents average change computed over ten runs of the algorithm. Performance of each feature group was characterized by comparing the AUC score of the forecasting algorithm once the feature group was added to the AUC score of the forecasting algorithm without the feature group. For example, in the case of physical activity, we compared the AUC score when the algorithm included all feature groups to the AUC score when the algorithm included only heart rate, sleep, and cycles feature groups.

### Held out evaluation cohort performance

The held-out evaluation cohort performed well in most cases (Figure 7 and Table 2). Performance (based on AUC scores) was better than chance performance in 7 of 8 (88%) people using the hourly forecaster (M = 0.68, SD = 0.18) and 4 of 8 (50%) people using the daily forecaster (M = 0.58, SD = 0.16). It is important to note that the participant, P7, who did not perform better than chance in the hourly forecast had the lowest seizure count during the evaluation period and was the only participant without a significant multiday cycle.

**Figure 7.**
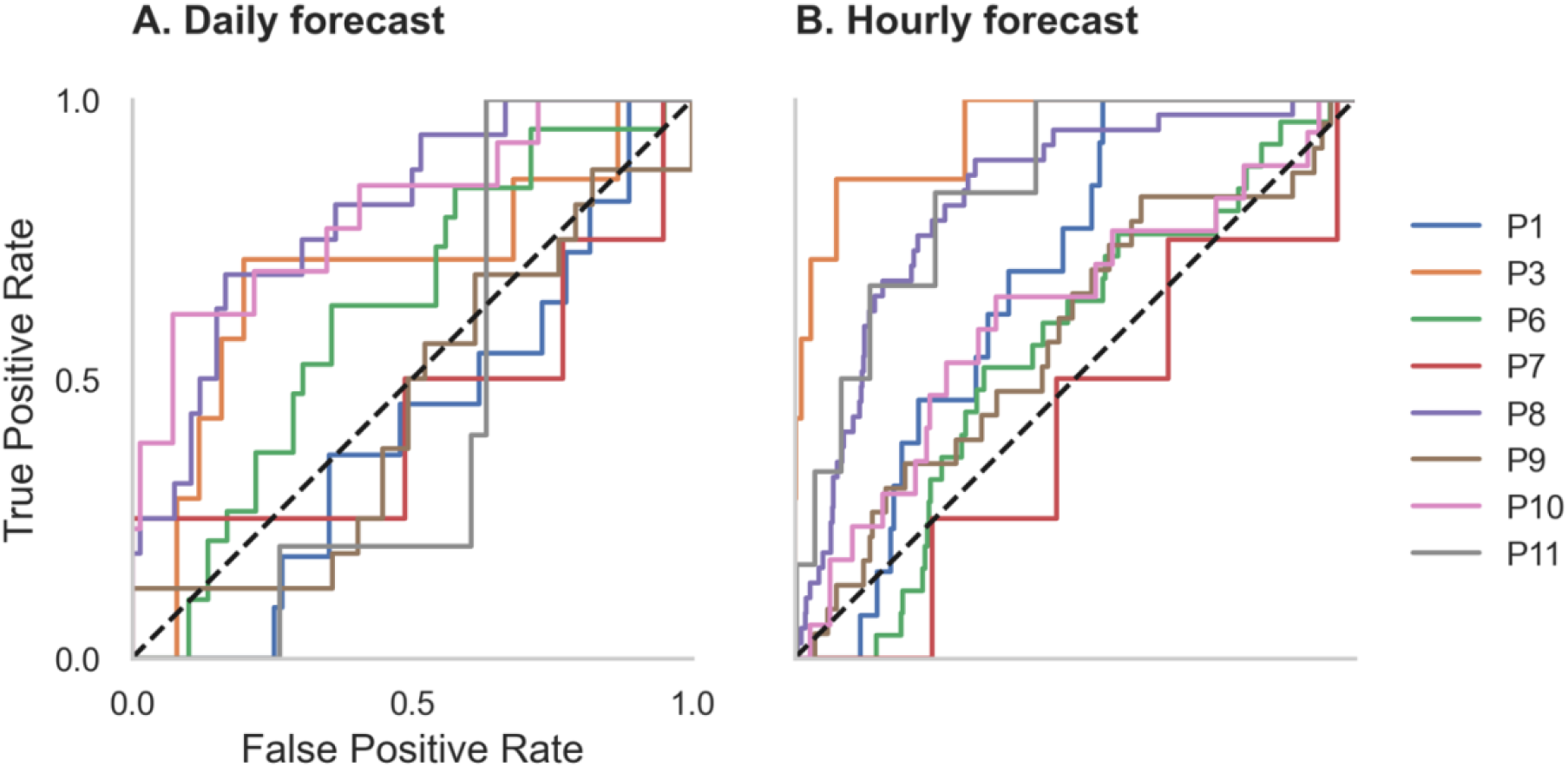
Receiver operator characteristic (ROC) curves for all participants in the daily and hourly forecast (held-out evaluation testing cohort). The dashed diagonal line represents a balanced random forecast. ROC curves show that hourly forecasts mostly outperformed a balanced random forecaster, and daily forecasts outperformed a balanced random forecaster half of the time. Patient-specific forecast performance was assessed by comparing the forecaster’s area under the ROC curve (AUC) to the AUC of a rate-matched random forecast (different to the balanced random forecast shown above).

## Discussion

### Summary

People with epilepsy and their caregivers have expressed their interest in non-invasive wearable devices for decades, particularly for seizure forecasting (44) and detection (45). Wearable devices are more acceptable to people with epilepsy than invasive, cumbersome or indiscrete devices (44,45). Nonetheless, very few studies have investigated the feasibility of non-invasive wearables in seizure forecasting.

This study demonstrates that features recorded via non-invasive wearable sensors can contribute to accurate seizure forecasts. Individual forecasters performed better than chance with all people when an hourly prediction horizon was used, and with 10 of 11 people when a daily prediction horizon was used. These results indicate that non-invasive seizure forecasting is possible for people with epilepsy with seizure warning periods of up to 24 hours.

In the evaluation cohort, forecasters performed better than chance in 7 of 8 people using the hourly forecaster and 4 of 8 people using the daily forecaster. This is contrary to what we expected: that the performance would improve with a longer period on which to train the algorithm. The lack of improvement in AUC scores in the evaluation cohort may be attributed to the shorter recording lengths and seizure counts in the evaluation dataset compared to the testing dataset, making it difficult to directly compare the cohorts. Furthermore, the theoretical shift and change that may occur in heart rate cycles over time was not considered in this model. This shift in cycles may be mitigated by consistently retraining the algorithm on a shorter period of data (e.g., the past four months, instead of all past data).

Generally, the hourly forecaster performed better than the daily forecaster. The superior performance in the hourly forecasts may be attributed to a number of factors, such as the inclusion of circadian heart rate cycles, hourly step count and RCH. The resolution of the daily forecaster would also have played a role in the loss of information. For example, high frequency seizure days (>1 seizure occurred on a day) were weighted equally to low seizure frequency days (1 seizure on a day).

### Feature importance

Overall, cyclic features (heart rate cycles and previous seizure timing) were the strongest predictors of seizures in most cases (Figure 6). However, sleep, physical activity, and other heart rate features were also valuable predictors in some cases.

Sleep features were useful predictors of seizure likelihood for some people using the daily forecaster but were weak predictors in the hourly forecaster. The lack of utility of sleep features in the hourly forecaster may be attributed to the design of the algorithm, as the sleep variable remains constant for all hours of the day after waking, making it difficult for the algorithm to distinguish between non-seizure and seizure hours. In contrast, sleep was a useful feature in the daily forecaster, which distinguishes seizure-days from non-seizure days, suggesting that sleep does play a role in seizure risk for some people. Sleep features, such as sleep quality, transitions and length, have historically been associated with seizures in many people with epilepsy (18,23). It is possible that the role of sleep as a seizure precipitant is highly patient-specific, which warrants further investigation in larger cohorts.

Heart rate features – daily RHR and RCH (estimation of HRV) – were useful in predicting seizure likelihood in some people (Figure 6). HRV has been of interest to researchers for decades and is known to reflect autonomic function (46). HRV has also been used to predict seizures minutes in advance, albeit with high false prediction rates (39). It is important to note, however, that we have only estimated HRV in this study, and further work should investigate whether heart rate variability can be accurately estimated from photoplethysmography. Daily resting heart rate, on the other hand, is not often associated with seizure risk, but seemed to be a useful feature in some cases. However, daily resting heart rate is likely correlated with multiday rhythms of heart rate and thus may not provide distinct value compared to cyclic features that were derived from heart rate.

Physical activity features were also predictive of seizures in some people, particularly in the hourly forecaster (Figure 6). Physical activity is beneficial for mental health, quality of life, and cognitive function for people with epilepsy (47). However, people with epilepsy are less likely to engage in physical activity than the general population (48), partially influenced by the inaccurate historical belief that exercise can provoke seizures (49). On the contrary, there is some evidence that increased physical activity is associated with reduced seizure frequency (50,51). Physical activity is also known to benefit common psychiatric comorbidities of epilepsy, such as anxiety and depression (52), so exercise may indirectly reduce seizure frequency by impacting other seizure precipitants, such as stress and reduced heart rate. We did not explore whether the relationship between physical activity was generally positive or negative in this study, but this should be investigated in future work.

### Demographic and clinical factors

We generally observed that participants with longer recording times performed better, and more consistently over prediction horizons (Supplementary Figure 1). This suggests that seizure forecasts utilizing wearable sensors perform better with longer recording times, and are likely to improve over time. We suggest that a clinical forecast requires a minimum amount of data or events before starting to use the forecaster. Future work should investigate the ideal number of events required for the best results, taking into account an individual’s seizure frequency, and the optimal number of cycles to observe before incorporating the cycle into the forecaster.

Interestingly, participants with lower seizure frequencies tended to perform better in the hourly and daily forecasters (Supplementary Figure 2). This relationship between seizure frequency and forecasting performance was also observed in a prospective forecasting study (16). Although it is well known that seizure frequency is important to quality of life (53), people with fewer seizures are still subject to anxiety and fear caused by the unpredictability of seizures (8,54). Therefore, people who have fewer seizures may have the most to benefit from accurate forecaster, as a forecaster may enable them to go about their daily lives without fear of an impending seizure.

Despite less than perfect accuracy in the current model, the results may still meet the user requirements for a practical seizure gauge device. Many people with epilepsy may use a forecasting device despite less than perfect accuracy (44). For example, subjects in a prospective seizure forecasting study found the implanted device useful even though the median sensitivity was only 60% (55). Moreover, shorter time horizons (minutes to hours) seem to be preferable over longer time horizons (days) (44). This is in line with the current results, where the shorter time horizon (hourly) performed better than the longer time horizon (daily). Ultimately, prospective seizure forecasting studies with non-invasive wearables are needed to assess user requirements and clinical utility.

### Limitations

This study has several limitations. First, self-reported seizure diaries have inherent drawbacks and are known to be inaccurate (56). Not everyone with epilepsy is aware of when they experience a seizure, particularly if they predominantly have focal aware seizures. However, self-reported events are non-invasive, easy to capture, and remain the standard data source for medical practice and clinical trials in epilepsy (10). Therefore, seizure diaries remain important for non-invasive seizure forecasting. To improve the accuracy of self-reported events, non-invasive seizure detection devices are available for convulsive seizures, and detection of nonconvulsive seizures are in the pipeline (57). Second, it is worth noting that the accuracy of heart rate and sleep stages measured from smartwatch devices has been investigated compared to electrocardiography and polysomnography, respectively (58–60). Smartwatches are known to be useful in obtaining gross estimates of sleep parameters and heart rate but are not yet suitable substitutes for electrocardiography and polysomnography. This suggests that complex parameters, such as sleep stages and heart rate variability, may need further investigation to understand their role as seizure drivers. Third, seizure number and seizure frequency are also limiting factors on whether seizure forecasting is possible. When seizure numbers are low, the forecaster may be unreliable in some cases due to overfitting in the training set. The optimal learning period based on seizure frequency should be investigated in future. Fourth, cyclic variables were the most dominant features in both the hourly and daily forecasters, but these cycles may shift or change over time, thus affecting the accuracy of the forecaster. In the current study, we tested the model using the same cycles derived during initial training of the algorithm. This should be considered in future analyses. Finally, we attempted to balance our participant recruitment so that it accurately reflected the population of people with refractory epilepsy (variety of adult ages, epilepsy types and seizure frequencies); however, the limited number of participants in this study means that the population may not have been accurately represented in the sample, particularly for people with generalized epilepsy. We also endeavor to explore the relationship between forecasting accuracy and epilepsy type in the future.

### Conclusion

We assessed the utility of electronic self-reported seizure diaries and non-invasive wearable physiological sensor data to estimate seizure risk in retrospective and pseudo-prospective cohorts. This research has shown that non-invasive wearable sensors in the field of seizure forecasting is not only possible, but feasible and imminent. Prospective analysis and clinical trials should also be undertaken on longitudinal datasets in the future.

## Data Availability

Code used in this manuscript is available upon request.

