## Supplementary Figure 1, Supplementary Figure 2 for "Forecasting seizure likelihood with wearable technology"

### Supplement

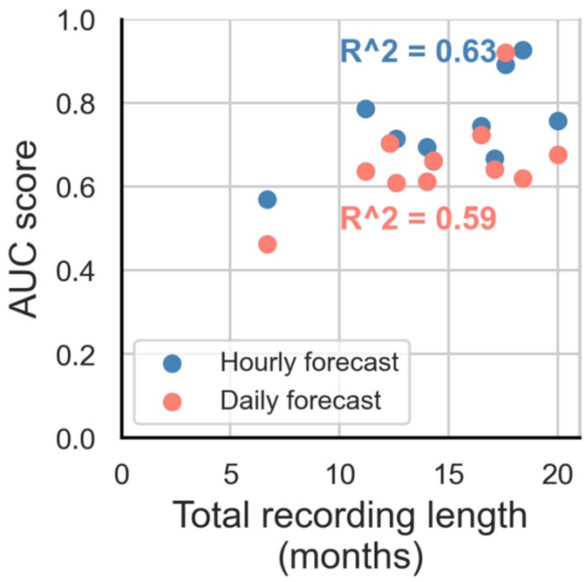


**Supplementary Figure 1.** The relationship between AUC scores (shown in Figure 2) and recording length (months), shown for all participants. AUC scores were shown for both the hourly forecaster (blue) and the daily forecaster (orange). The pearson correlation coefficient ($R^2$) of each forecaster is also shown.

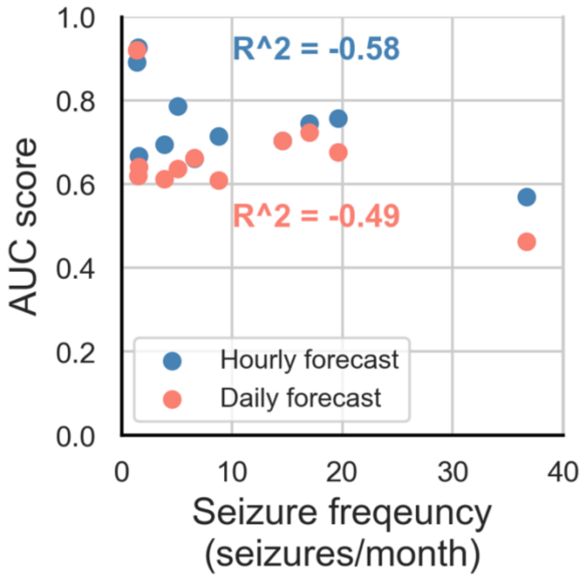


**Supplementary Figure 2.** The relationship between AUC scores (shown in Figure 2) and seizure frequency (seizures/months), shown for all participants. AUC scores were shown for both the hourly forecaster (blue) and the daily forecaster (orange). The pearson correlation coefficient ($R^2$) of each forecaster is also shown.
